# Assessing the impact of dental and periodontal statuses on the salivary microbiome: a global oral health scale

**DOI:** 10.1101/2020.05.16.20104190

**Authors:** M Relvas, A Regueira-Iglesias, C Balsa-Castro, F Salazar, JJ Pacheco, C Cabral, C Henriques, I Tomás

**Affiliations:** Institute of Research and Advanced Training in Health Sciences and Tecnologies (IINFACTS), IUCS-Cespu-Instituto Universitário de Ciencias da Saúde, Gandra, Paredes, Portugal; Oral Sciences Research Group, Special Needs Unit, Department of Surgery and Medical-surgical Specialties, School of Medicine and Dentistry, Health Research Institute of Santiago (IDIS), Universidade de Santiago de Compostela, Galicia, Spain

**Keywords:** supragingival plaque, caries, periodontitis, 16S rRNA gene, salivary microbiome, core species, differential abundance

## Abstract

Very few 16S rRNA-based studies have conducted a simultaneous analysis to identify the impact of various dental and periodontal parameters and determine which of them have the greatest repercussion for the salivary microbiota. Consequently, this study used 16S rRNA gene amplicon sequencing to assess the impact on salivary microbiome of different grades of dental, periodontal and global oral disease.

Our global oral health scale was used to produce a convenience sample of 81 patients from 270 who were initially recruited. These subjects were assigned the following grades: 47 had a periodontal grade (PG) of 0 and dental grades (DGs) between 0-3, and 46 had a DG of 0 and PGs between 0-3. Saliva samples were collected from each participant. Sequencing was performed in Illumina MiSeq with 2 × 300 bp reads, while the raw reads were processed according to the Mothur pipeline. The statistical analysis of the 16S rDNA sequencing data at the species level was conducted using the Phyloseq, DESeq2 and Microbiome packages. The impact on the salivary microbiota of the different DGs, PGs and global oral grades (GGs) was investigated in relation to: 1) indicators of alpha diversity and the structure of the bacterial community; and 2) the composition of the core microbiome and the results of differential abundance tests.

The simultaneous presence of dental and periodontal pathology has a potentiating effect on the richness and diversity of the salivary microbiota. The structure of the bacterial community in oral health differs from that present in dental, periodontal or global oral disease, especially in high grades. The non-specific microbiome core contains a greater number of more abundant species than the specific core of a particular dental or periodontal condition (health or pathology). The number of taxa in the salivary microbiota with differential abundances between the DGs, PGs or GGs represents, at most, a quarter of the bacterial community and are mainly non-core species. Supragingival dental parameters influence the microbiota’s abundance more than subgingival periodontal parameters, with the former making a greater contribution to the impact that global oral health has on salivary microbiome.

## INTRODUCTION

Oral diseases are a major public health concern, having a negative effect on individuals, communities and society at large (1). Recent data from the Global Burden of Disease Study (GBD) revealed that around 3.5 billion people worldwide have suffered from an untreated dental condition, mainly in the form of caries, periodontitis, tooth loss and edentulism (2). It is also well-known that oral diseases such as periodontitis have significant associations with chronic systemic conditions like cardiovascular disease and diabetes, with poor oral health having an adverse impact on the development and control of these types of disorder (3-5).

Several sets of diagnostic criteria have been developed over the last few decades for use in the classification of dental caries. These include: the Decayed, Missing and Filled Teeth (DMFT) index (6); the Dundee Selectable Threshold Method (DSTM) (7); the International Caries Detection and Assessment System (ICDAS) (8); and the Caries Assessment Spectrum and Treatment (CAST) tool (9). The Community Periodontal Index of Treatment Needs (CPITN) (10) and the recent International Classification of Periodontal and Peri-implant Diseases and Conditions are also used to categorise periodontal status, with the latter distinguishing forms of the disease based on a multidimensional staging and grading system (11,12).

In recent years, scales have been designed based on a combination of different dental and/or periodontal parameters. These include: the Total Dental Index (TDI) (13); the Modified Total Dental Index (MTDI) (14, 15); the Dental Asymptotic Score (DAS) (16); and the Brief Oral Health Status Examination (BOHSE) (17). In 2013, our group developed a new (adult population) global oral health scale, combining parameters relating to caries, gingivitis and periodontitis. The aim of this clinical scale, which is applicable to the adult population, is to obtain a single numerical value that synthesises the patient’s oral health status, allowing the establishment of an individualised grade of dental or periodontal health (18).

A symbiotic relationship between resident oral microbiota and a host is essential for the maintenance of oral health (19, 20). Indeed, several studies using 16S rRNA gene next-generation sequencing (NGS) have shown that local compositional changes in the bacterial communities, which are associated with dental caries and periodontitis, are far more complex than previously believed (21,22). Assuming saliva may reflect the presence of oral disease (23), investigations using 16S rRNA gene NGS have been conducted to identify which salivary microbiota profiles are related to oral health (24), dental caries (25) and periodontitis (26).

However, very few 16S-based studies have analysed the impact of various dental and periodontal parameters simultaneously to demonstrate which of them have the greatest repercussions for the salivary microbiota (27-29). As a consequence, our study used 16S rRNA gene NGS to examine the bacterial diversity of the salivary microbiota in patients who have been classified clinically by our self-designed and previously validated global oral health scale. Our main goal was to analyse the individualised impact of different grades of dental, periodontal and global oral disease on the salivary microbiota, as well as the extent to which this bacterial community is affected.

## MATERIALS AND METHODS

### Selection of the study group

The participants in our study were recruited between January, 2016 and July, 2017 from a group of patients aged between 25 and 65 who had visited the Dental Clinic of Instituto Superior de Ciências da Saúde Norte, Cooperativa de Ensino Superior, Politécnico e Universitário (CESPU) (Gandra, Paredes, Portugal) for a dental check-up.

The following exclusion criteria were applied: 1) fewer than 18 teeth; 2) the presence of removable dental prostheses or orthodontic devices; 3) antibiotic treatment or the routine use of oral antiseptics in the previous three months; and 4) the incidence of any etiological factors (systemic diseases, medication, radiotherapy etc.) that could trigger changes in the production and/or composition of saliva (30). After applying these criteria, 270 patients remained in our sample. The research was conducted based on the principles of the Declaration of Helsinki (revised in 2000) concerning studies involving human experimentation (31). The research protocol was approved by the Instituto Superior de Ciências da Saúde –Norte, CESPU (registration number 35/CE-IUCS/2019), and all the participants provided their written informed consent to their involvement in the study.

### Application of our oral health scale

Examination of the oral cavity (excluding the third molars) was performed by a single trained examiner using a conventional dental mirror, a probe, and a CP11 manual periodontal probe calibrated at 3, 6, 8 and 11 mm. Six sites per tooth were examined, taking into account the dental and periodontal variables included in our new scale. The findings are expressed in terms of the number of sites revealing specific oral conditions, and were evaluated according to tested and reproducible indices.

The following four clinical parameters were utilised: 1) the number of sites with supragingival plaque, based on the O’Leary Plaque Index (32); 2) the number of sites with caries and their severity (1= affecting the enamel, 2= affecting the enamel and dentine, and 3= affecting the enamel, dentine and pulp); 3) the number of sites with gingival inflammation, based on the Ainamo and Bay Gingival Index (33); and 4) the number of sites with periodontal pockets ≥4 mm and their severity (average depth of the pockets). The participants’ dental and periodontal health grades (DG and PG, respectively) corresponded to the grades assigned to at least two of the three variables analysed in each of these categories. If there were differences between the grades allocated to each of the variables in a category, the parameters for “number of caries” and “number of periodontal pockets ≥4 mm” took precedence. If the same grade was allocated to two variables in a category, but the third variable’s grade was two levels higher, the value assigned to it was one grade higher than that of the matching variables. The global oral health grade (global grade -GG-) was determined by the category (dental or periodontal) with the highest ranking, enabling patients to be classified based on the score for their dental, periodontal and global oral health (Table 1).

**Table 1.** Scale of global oral health.

| **Grades of Dental Health** | **Grade 0** | **Grade 1** | **Grade 2** | **Grade 3** |
| --- | --- | --- | --- | --- |
| Supragingival plaque (O'Leary index)* | 0 | 1-56 | 57-112 | >112 |
| Caries* | 0 | 1-4 | 5-8 | ≥9 |
| Severity of the caries (median) | 0 | 1 | 2 | 3 |
| **Grades of Periodontal Health** | **Grade 0** | **Grade 1** | **Grade 2** | **Grade 3** |
| Gingival inflammation (Ainamo and Bay index)* | 0 | 1-56 | 57-112 | >112 |
| Periodontal pockets ≥4 mm* | 0 | 1-56 | 57-112 | >112 |
| Severity of the pockets (mean) | <4 mm | 4-4.9 mm | 5-5.9 mm | ≥6 mm |

\*The determination of these parameters was done at the site level (six sites per tooth). The global oral health grade was determined by the category (dental or periodontal) with the highest ranking.

Our global oral health scale was used to select a convenience sample of 81 patients from the 270 initially recruited. These 81 participants were given the following GGs: 0 for 17 of them; 1 for 25; 2 for 28; and 3 for 11. In relation to the subscales, 47 patients had a PG of 0 and different DGs, and 46 had a DG of 0 and different PGs.

### Collection of saliva samples and the 16S rDNA gene amplicon sequencing

Unstimulated saliva samples (2–3 mL) were collected from each participant using the spitting method (34). None of the subjects had eaten or brushed their teeth for at least one hour before they provided their sample.

Total DNA was extracted from the saliva using a commercial kit (QIAamp DNA Mini kit; Qiagen, GmbH, Hilden, Germany) and according to the manufacturer’s instructions, albeit with minor modifications, including the addition of a lysozyme treatment (20 mg/ml at 37 °C for 30 min). The isolated DNA was eluted in 200 μl of distilled and apyrogenic water, and its quality and concentration were assessed using a Nanodrop spectrophotometer (ND-2000 Spectrophotometer, Wilmington, USA). DNA samples with spectrophotometer ratios (Abs 260/280) between 1.5 and 2.0 were considered to be acceptable for inclusion in the study (35). We used the ZymoBIOMICS™ Microbial Community DNA Standard (Catalog No. D6306, Zymo Research, Irvine, CA, USA), which is a mix of genomic DNA isolated from pure cultures of eight bacterial and two fungal strains, as both the bacterial mock community and the positive control for the downstream procedures. Mock DNAs were amplified and sequenced in the same way as all the other samples used in the experiment.

A polymerase chain reaction (PCR) amplification of the 16S rDNA gene was performed with the KAPA HiFi HotStart DNA Polymerase KAPA HiFi HotStart ReadyMixPCR Kit (Cat. No. KK2602, 7958935001; Kapa Biosystems, F. Hoffmann-La Roche Ltd, Basel, Switzerland). The V3-V4 hypervariable region was amplified as previously described (36) using the following universal primers in a limited-cycle PCR:

V3-V4-Forward (5’ - TCGTCGGCAGCGTCAGATGTGTATAAGAG ACAGCCTACGGGNGGCWGCAG-3)
V3-V4-Reverse (5’ - GTCTCGTGGGCTCGGAGATGTGTATAAGA GACAGGACTACHVGGGTATCTAATCC-3)

A set of modified primers, V3-V4-F and V3-V4-R, were also used. This set contained a 1-3 bp “heterogeneity spacer” that we designed to mitigate the issues caused by low sequence diversity amplicons.

Each PCR amplification was carried out on a total volume of 10 μl, which comprised 4 μl of DNA, 0.2-μM from each forward and reverse primer, and a Kapa ready mix (Kapa Biosystems). The PCR conditions were modified by conducting: 1) an initial denaturation at 95°C for 3 min; 2) 25 three-step cycles of 95°C for 30 s, 55°C for 30 s and 72°C for 30 s; and 3) a final 5 min extension at 72°C. Water, up to a total volume of 50 μl, was added after the first PCR step. The reactions were purified using AMPure XP beads (Beckman Coulter, Brea, CA, USA) with a 0.9X (V3-V4 amplicon) ratio, according to the manufacturer’s instructions. PCR products were eluted from the magnetic beads with 32 μl of Buffer EB (Qiagen N.V, Hilden; Germany), with 30 μl of the eluate transferred to a fresh 96-well plate. The primers described above contain overhangs that enable the addition of full-length Nextera adapters. Barcodes are available for multiplex sequencing in a second PCR step, which produces sequencing-ready libraries. To this end, 5 μl of the first amplification was used as a template for the second PCR, with Nextera XT v2 adaptor primers added up to a final volume of 50 μl. The PCR mix and thermal profile employed for the first PCR were also used for the second, but only for eight cycles. After the second PCR, 25 μl of the final product was purified and normalised with the SequalPrep normalisation kit (Invitrogen, Carlsbad, CA, USA), according to the manufacturer’s protocol. Libraries were eluted in a 20-μl volume and pooled for sequencing.

Final pools were quantified with a qPCR using the Kapa library quantification kit for Illumina Platforms (Kapa Biosystems) on an ABI 7900HT real-time cycler (Applied Biosystems, Foster City, CA, USA). Sequencing using v3 chemistry with a loading concentration of 18 pM was performed in Illumina MiSeq (Illumina Inc., San Diego, CA, USA) with 2 × 300 bp reads. In all cases, 10% of the PhIX control libraries were spiked to increase the diversity of the sequenced samples.

In parallel, negative control tests of the sample-collection buffer, DNA-extraction and PCR-amplification steps were conducted routinely under the same conditions and using reagents. One such non-template control was subjected to the library preparation and then sequenced. As expected, this yielded very few reads (220 per sample), in contrast to an average of 188,713 reads/library in the sample-derived collections. The sequences obtained were deposited in the SRA database under accession number PRJNA623352.

### Bioinformatic processing

The raw reads were obtained in the fastq format and processed according to the Mothur pipeline proposed by Schloss et al. (37), albeit with some modifications. Four samples with a very low number of raw sequences were removed (fewer than 436 per sample). The final raw-read total was 15,285,797 (mean ± standard deviation per sample = 188713 ± 37473, median = 189918, maximum-minimum number of sequences in a sample = 282936-119882).

The Needleman-Wunsch alignment algorithm was used to obtain the contigs from the raw sequences (38). The quality filtering criteria applied were contigs with: 1) errors in the primers; 2) ambiguous bases; 3) more than eight homopolymers; 4) mismatches; and 5) overlap of less than 25 bp. Lengths below 400 bp or greater than 535 bp were discarded.

The unique candidate sequences were aligned against the SILVA-based bacterial reference alignment (39), again using the Needleman-Wunsch algorithm (38). A total of 52,544 potential chimeras were detected with the VSEARCH tool (40), representing 4.06% of the total unique sequences. The final total of high-quality reads was 4,269,754.

The opticlust algorithm and the Matthews Correlation Coefficient Metric (MCC) were used to group the sequences into operational taxonomic units (OTUs) with a similarity of 97% (41). A total of 3,788 OTUs were obtained from all the samples, and the native Bayesian classifier proposed by Wang et al. (42) was employed to categorise them. The Human Oral Microbiome Database, version 15.2, was used for this purpose (43) and includes 1015 different consensus strains. A classification threshold of 80% was employed for the taxonomic designations at all levels. The non-classified level was assigned the term “unclassified.”

The mock D6306 sample was processed using the same protocol to evaluate the quality of the sequencing and the bioinformatic pipeline. First, a reference database was constructed from the 58 different 16s rRNA sequences provided by the manufacturer. These corresponded to all the variants of the 16s rRNA genes of the eight species included in the sample. The mock sample was then extracted from the pipeline and analysed separately against the reference database, producing errors of 0.0000073% in the bp from a total of 57,350 sequences with a length of 400 bp. Fourteen OTUs were obtained from the mock sample, with eight of them highly abundant (between 4241 and 14971 counts) and present in very similar numbers to the theoretical percentages indicated by the manufacturer. Only six OTUs were present in very low abundances (one or two counts), which could be due to sequencing errors. As a consequence, the protocol applied was considered to be of high quality.

### Statistical analysis

The statistical analysis of the 16S rDNA sequencing data at the species level was performed according to the protocol recently proposed by McMurdie and Holmes (44), using implementations in R such as the Phyloseq, DESeq2 and Microbiome packages (45-47). An independent filter had previously excluded from the statistical analysis the OTUs with an abundance of ≤2 counts and a presence in ≤3 samples (3.89%) (48), leaving a final total of 403 OTUs(the final number of high-quality reads was 4,235,218.

The impact of the different oral-health scale grades (DG, PG and GG) on the salivary microbiome was investigated in relation to: 1) four indicators of alpha diversity and the structure of the bacterial community; and 2) the composition of the core microbiota and the testing of differential abundances. The individualised impact of the dental subscale was analysed in the participants with a PG of 0 while the periodontal subscale was assessed in those with a DG of 0. Four different comparative analyses of the grades for the dental and periodontal subscales and global oral health scale were performed: grade 0 vs 123; grade 0 vs 1; grade 0 vs 23; and grade 1 vs 23.

#### Alpha diversity indicators and structure of the salivary bacteria community

The phyloseq and microbiome packages were used in the following ways to obtain data relating to the four alpha diversity indicators (45,47): 1) taxa richness: the absolute count data (“observed”) and the coverage index; the latter specifies how many of the more abundant OTUs are required to achieve a particular proportion of the occupied ecosystem (95%); and 2) diversity and evenness of the taxa present in the samples: the Shannon and Pielou indices (49,50). The Student t-test was used to conduct different comparative analyses.

A principal coordinate analysis (PCoA) was employed to visualise the clustering of the salivary samples in relation to their respective global, dental and periodontal health grades. The “ape” package, version 5.3 (51), was used to rotate the phylogenetic tree, while the weighted unifrac algorithm (52) was applied in the phyloseq package to obtain the phylogenetic distance matrix. A non-parametric permutational multivariate analysis of variance (PERMANOVA) (53) was used to measure the multivariate community-level differences between the groups. These analyses were performed using the vegan package, version 2.5.6 (54).

#### Composition of the core salivary microbiome and testing the differential abundances

We used the microbiome package (47) to identify the central taxa present with a prevalence rate of 100% for each grade, or combination of grades from the dental and periodontal subscales and the global scale.

The DESeq2 package (46) was used to identify the bacteria with the most significant changes in differential abundance at the species level for the different conditions. Improvements to the stability and dispersion of the counts (variance) were required before it was possible to calculate the differential abundances for the different species present in the samples being compared. To this end, we used the estimateSizeFactors function in DESeq2 (46) to transform the stabilisation of the variance. The differential abundances were measured with the log2foldchange value, and the different conditions were compared using the Wald test with the Benjamini-Hochberg correction (Q parameter = 0.1, FDR < 10%). The differential-abundance measurements were statistically significant if the adjusted p-value was <0.05 (-log10 adjusted p-value =1.3). Figure 1 shows the study’s methodological protocol, in particular concerning the participants’ clinical evaluations, the collection of the saliva samples, bioinformatic processing and the statistical analysis of the 16 S rRNA gene sequencing data.

**Figure 1.**
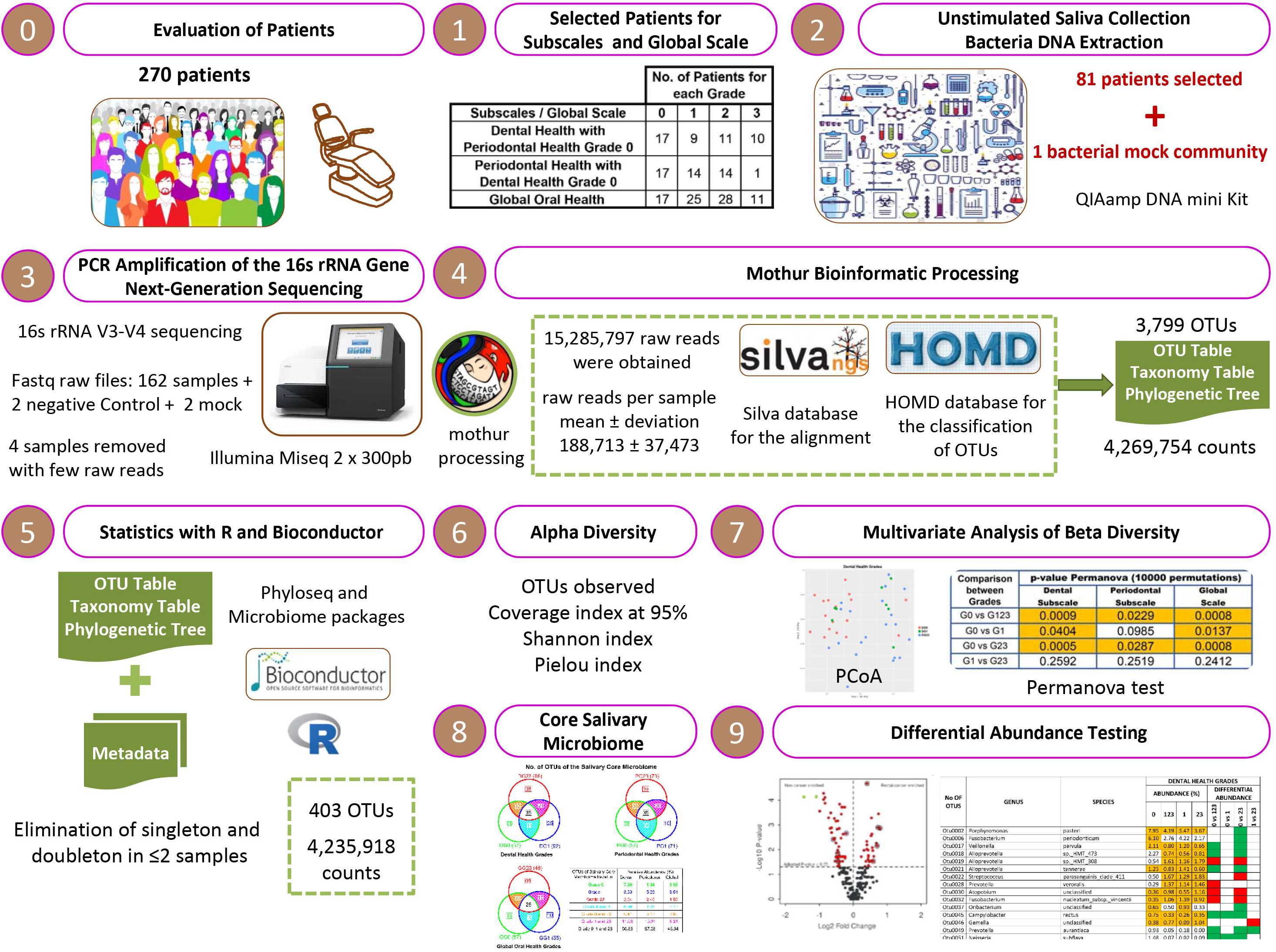
The methodological protocol used in the present study.

## RESULTS

The 81 participants in the convenience sample had the following GG scores: 0 for 17 of them; 1 for 25; 2 for 28; and 3 for 11. In relation to the subscales, the results were as follows: 1) 47 patients had a PG of 0 and DGs between 0 and 3 (nine had a DG of 1, 11 a DG of 2, and 10 a DG of 3); and 2) 46 had a DG of 0 and PGs between 0 and 3 (14 had a PG of 1; 14 a PG of 2; and one a PG of 3). The four patients excluded due to the low number of raw sequences obtained were: two of GG1 and two of GG2 (two of DG0, one of DG1 and one of DG2; two of PG0, one of PG1 and one of PG2).

### Impact of the dental and periodontal subscales and the global oral health scale on the salivary microbiome: alpha diversity indicators and the structure of the bacterial community

As seen in Table 2, worsening dental or periodontal health revealed a trend of increasing alpha diversity in both the dental and periodontal subscales. In our comparisons, however, significant differences in the number of OTUs were only observed in the former: DG0 *vs* DG123 (209.82 ± 37.01 vs 240.57 ± 35.02; p= 0.009); and DG0 vs DG23 (209.82 ± 37.01 *vs* 243.55 ± 33.09, p= 0.006). Meanwhile, in our global oral health scale comparisons, grade increments were linked to progressive increases in bacterial richness, especially in GG0 vs GG23 (no. observed OTUs= 212.52 ± 38.44 vs 240.24 ± 32.77, p= 0.015; coverage index 95%= 65.17 ± 15.59 *vs* 78.24 ± 20.42, p= 0.013). This was also the case for the Shannon Index values (GG0= 3.67 ± 0.23, and GG23= 3.83 ± 0.26, p= 0.026).

**Table 2.** Indicators of alpha diversity at different grades of the dental and periodontal subscales and the global oral health scale.

| Dental Health Grade (DG) | No. Observed OTUs | Coverage index (95%) | Shannon index | Pielou index |
| --- | --- | --- | --- | --- |
| DG0 | 209.82 ± 37.01 | 65.11 ± 15.64 | 3.66 ± 0.23 | 0.68 ± 0.03 |
| DG123 | 240.57 ± 35.02 | 74.64 ± 20.54 | 3.79 ± 0.24 | 0.69 ± 0.03 |
| DG1 | 233.12 ± 40.84 | 71.37 ± 20.97 | 3.79 ± 0.26 | 0.69 ± 0.04 |
| DG23 | 243.55 ± 33.09 | 75.95 ± 20.76 | 3.79 ± 0.23 | 0.69 ± 0.03 |
| Significant Grade Comparisons* (p-value <0.05) | 0 vs 123<br>-<br>0 vs 23<br>- | -<br>-<br>-<br>- | -<br>-<br>-<br>- | -<br>-<br>-<br>- |
| Periodontal Health Grade (PG) | No. Observed OTUs | Coverage index (95%) | Shannon index | Pielou index |
| PG0 | 208.52 ± 35.75 | 64.88 ± 15.36 | 3.66 ± 0.23 | 0.68 ± 0.03 |
| PG123 | 219.48 ± 32.32 | 71.51 ± 18.80 | 3.76 ± 0.27 | 0.70 ± 0.04 |
| PG1 | 214.61 ± 39.62 | 66.07 ± 18.30 | 3.69 ± 0.24 | 0.68 ± 0.03 |
| PG23 | 224.00 ± 24.38 | 76.57 ± 18.45 | 3.83 ± 0.28 | 0.70 ± 0.04 |
| Significant Grade Comparisons* (p-value <0.05) | -<br>-<br>-<br>- | -<br>-<br>-<br>- | -<br>-<br>-<br>- | -<br>-<br>-<br>- |
| Global Oral Health Grade (GG) | No. Observed OTUs | Coverage index (95%) | Shannon index | Pielou index |
| GG0 | 212.52 ± 38.44 | 65.17 ± 15.59 | 3.67 ± 0.23 | 0.68 ± 0.03 |
| GG123 | 234.85 ± 36.47 | 75.28 ± 20.39 | 3.80 ± 0.26 | 0.69 ± 0.04 |
| GG1 | 226.17 ± 41.02 | 70.52 ± 19.86 | 3.76 ± 0.26 | 0.69 ± 0.03 |
| GG23 | 240.24 ± 32.77 | 78.24 ± 20.42 | 3.83 ± 0.26 | 0.70 ± 0.04 |
| Significant Grade Comparisons* (p-value <0.05) | 0 vs 123<br>-<br>0 vs 23<br>- | 0 vs 123<br>-<br>0 vs 23<br>- | 0 vs 123<br>-<br>0 vs 23<br>- | -<br>-<br>-<br>- |

The following four comparisons were performed: grade 0 vs grade 123; grade 0 vs grade 1; grade 0 vs grade 23; grade 1 vs grade 23.

The PCoA revealed a grouping of the salivary samples taken from the participants with DGs, PGs and GGs of 0.0. This was in contrast to the picture for the other grades, whose compositional distributions were more diverse (Figure 2). This visual observation was confirmed by the PERMANOVA test, which produced significant results for the comparison of grades 0 and 123 (dental subscale, p= 0.0009; periodontal subscale, p= 0.0229; global scale, p= 0.0008). These findings were mainly at the expense of the contrast between grades 0 and 23 (dental subscale, p= 0.0005; periodontal subscale, p= 0.0287; global scale, p= 0.0008).

**Figure 2.**
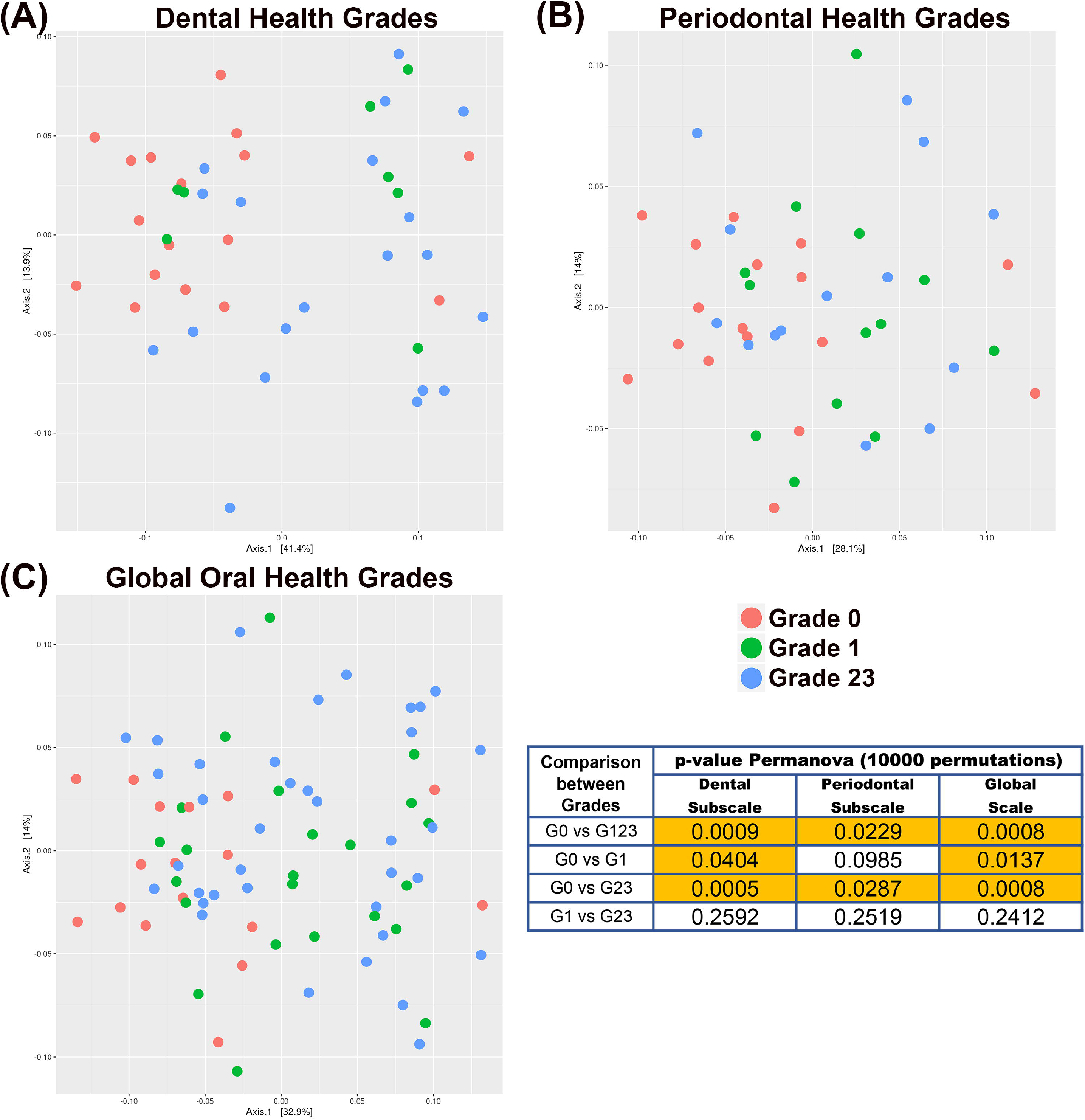
Principal Coordinate Analysis (PCoA), including PERMANOVA test values in the comparison between different grades of dental, periodontal and global oral health.

### Impact of the dental and periodontal subscales and the global oral health scale on the salivary microbiome: composition of the core microbiome and testing differential abundance

#### Composition of the core microbiome

Figure 3 uses Venn diagrams to portray the core microbiome’s number of OTUs for the different DGs, PGs and GGs. The core microbiome associated with the participants’ dental and periodontal health contained 57 species, representing 14.14% of the total number of OTUs and 63.06% of the total abundance. There were only nine taxa in DG0 and eight in PG0 (the specific core of grade 0), exemplifying abundances of 7.80% and 1.34%, respectively. Of these specific core species, five were common to both the dental and periodontal health conditions: *Neisseria macacae, Butyrivibrio* sp. *HMT_455, Campylobacter concisus, Porphyromonas catoniae* and *Corynebacterium durum*.

**Figure 3.**
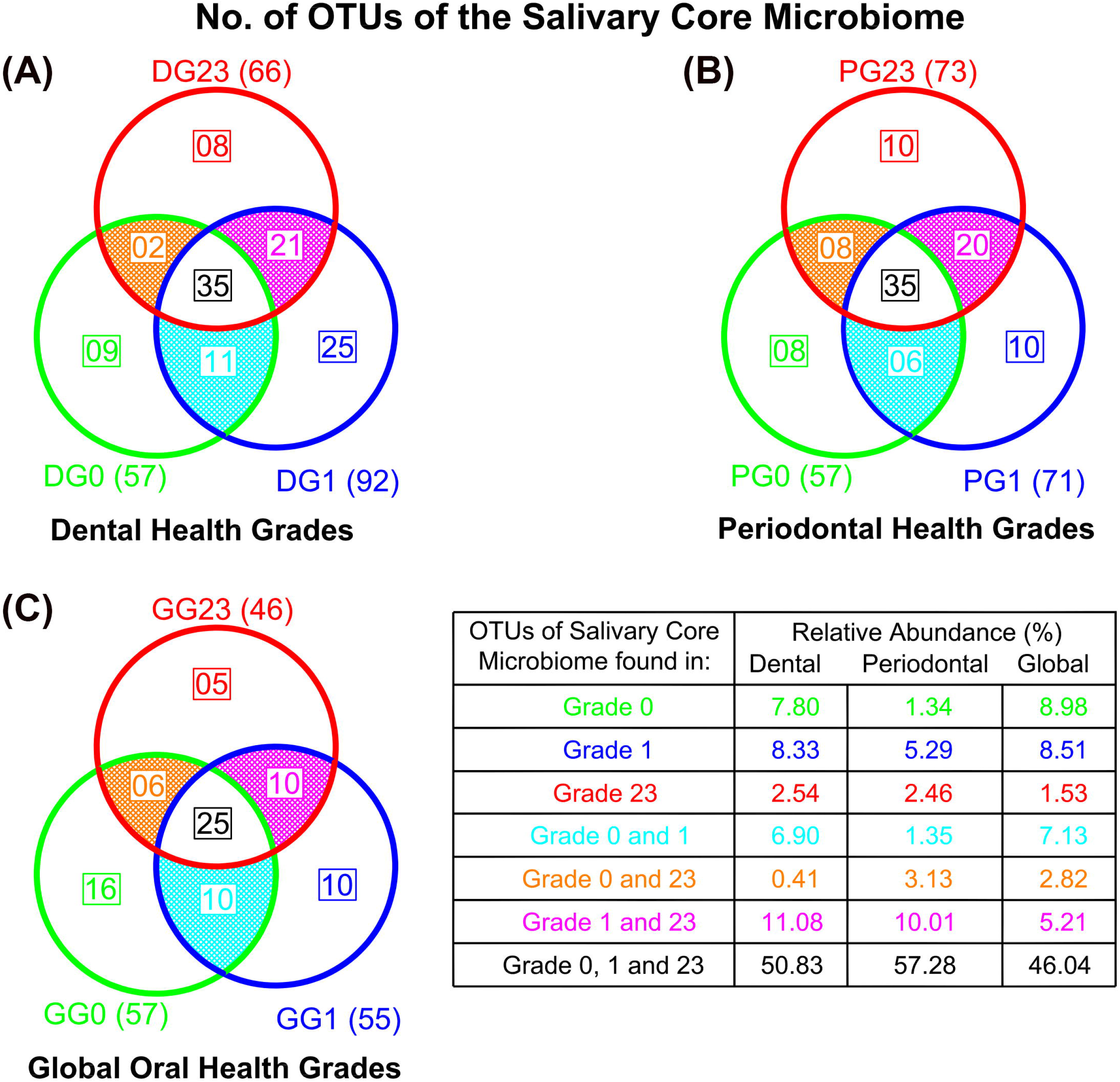
Number of OTUs of the salivary core microbiome present in different grades of dental, periodontal and global oral health, as well as their respective relative abundance values.

There were 66 species in the core microbiome associated with the most severe dental disease (DG23) and 73 with the most serious periodontal disease (PG23), representing 16.37% and 18.11% of the total salivary microbiota, and 67.14% and 67.98% of the total abundance, respectively. There were only eight and 10 taxa present in GG23 and PG23 (the specific core of grade 23), exemplifying 2.54% and 2.46% of the abundance, respectively. Of these specific core species, only *Porphyromonas endodontalis* was common to both pathological conditions.

There were 35 taxa common to both the dental and periodontal subscales, regardless of the grade (non-specific core), representing abundances of 50.83% and 52.75%, respectively. Of these non-specific core species, 25 were common to both subscales, with the most abundant being (abundance >1%): *Granulicatella adiacens, Haemophilus parainfluenzae, Leptotrichia* sp., *Porphyromonas pasteri, Prevotella* sp., *Prevotella melaninogenica, Prevotella salivae, Rothia mucilaginosa, Streptococcus* sp., *Streptococcus. oralis_subsp._dentisani_clade_058, Streptococcus salivarius, Veillonella* sp. and *Veillonella parvula*. Species belonging to the salivary core microbiome are shown in the supplementary data 1.

#### Testing differential abundance

The results for the dental health subscale revealed differential abundances for the different grades in 102 species (25.31% of the total OTUs), 39 of which were core species and 63 non-core (9.67% and 15.63% of the total OTUs, respectively). There were 36 species associated with oral health and 66 with some grade of dental pathology. In the comparison of DG0 vs DG23, there were differential abundances in the main dental-health related OTUs (with >1% levels in DG0; log2foldchange values ranged from 5.86 to 1.41): *Porphyromonas pasteri, Fusobacterium periodonticum, Veillonella parvula, Alloprevotella* sp. *HMT 473, Alloprevotella tannerae* and *Neisseria subflava*. Those related to high grades of dental pathology (>0.5% levels in DG23; log2foldchange values ranged from −1.98 to −1.35) were: *Alloprevotella* sp. *HMT 308, Streptococcus parasanguinis* clade *411, Atopobium* sp., *Fusobacterium nucleatum_subsp._vincentii, Megasphaera micronuciformis* and *Alloprevotella rava* (Table 3).

**Table 3.**
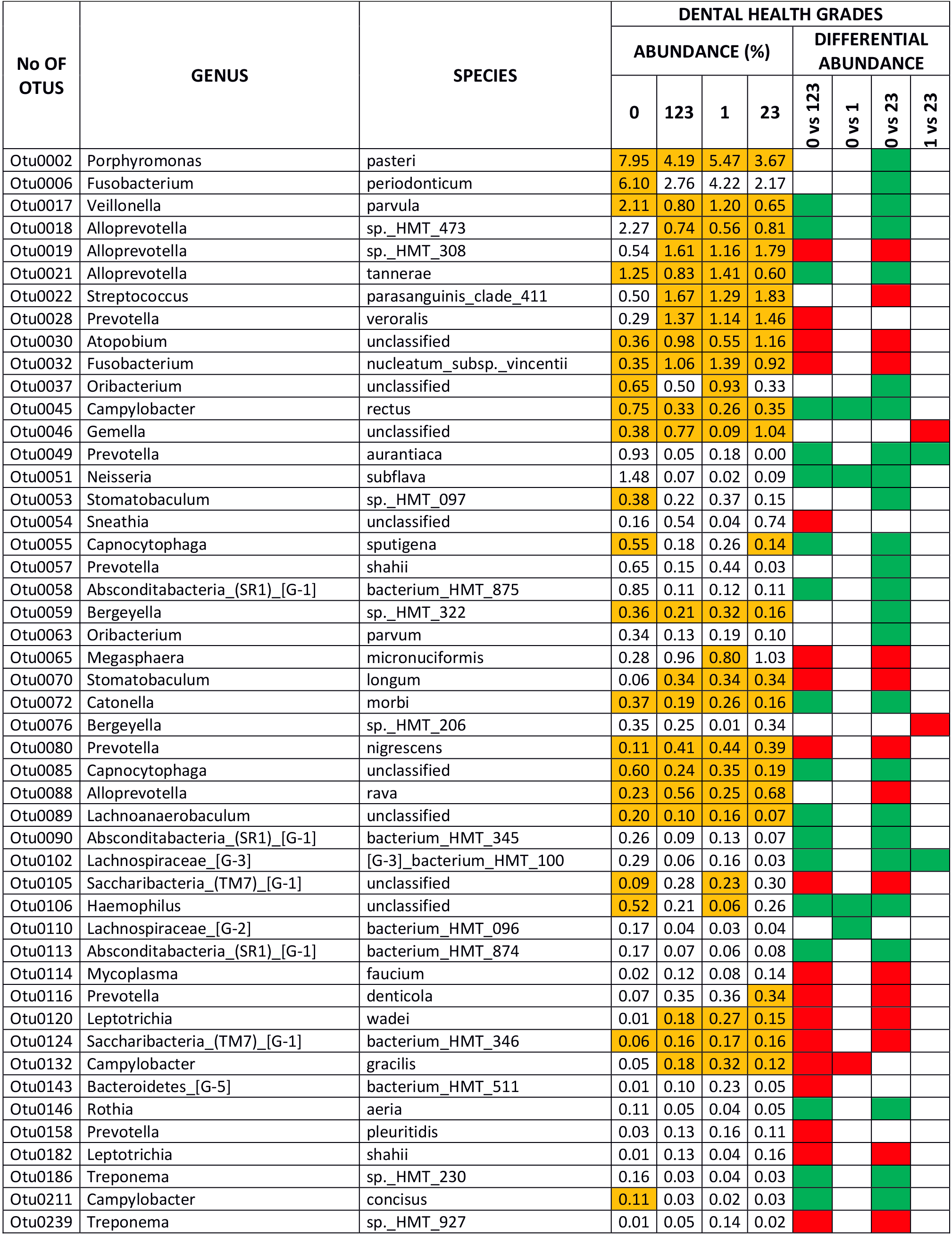
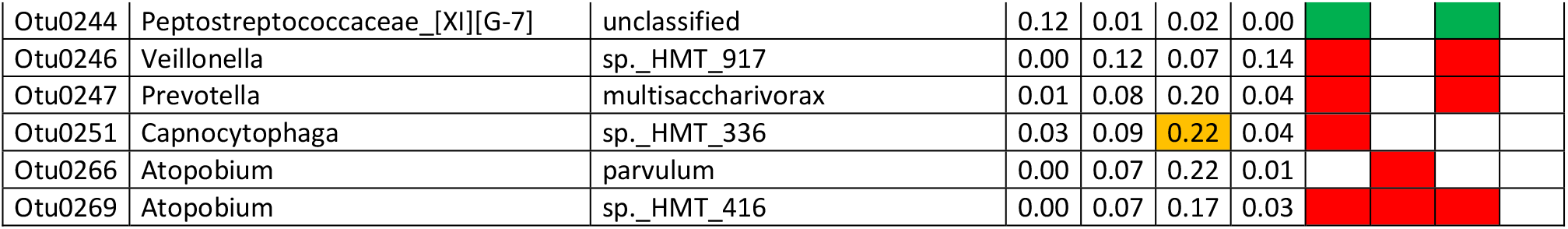
Species with abundance ≥0.1% at some grade and that presented differential abundance between the different grades of dental health (green square in favour of health; red square in favour of pathology). The species that belonged to the core of a certain grade are coloured orange.

The results for the periodontal health subscale revealed differential abundances for the different grades in 27 species (6.69% of the total OTUs), eight of which were core species and 19 non-core (1.98% and 4.71% of the total OTUs, respectively). Twelve species were associated with oral health and 15 with some grade of dental pathology. In the comparison of PG0 vs PG23, there were differential abundances in the two main periodontal-health related OTUs (with >1% levels in GG0): *Haemophilus parainfluenzae* and *Capnocytophaga leadbetteri*, with log2foldchange values of 1.16 and 2.31, respectively. Those associated with high grades of periodontal pathology (with >0.1% levels in GG23; log2foldchange values ranged from −3.07 to −2.13) were: *Tannerella forsythia*, *Mycoplasma faucium, Fretibacterium* sp., and *Bacteroidetes [G-5]* bacterium *HMT 511* (Table 4).

**Table 4.** Species that presented differential abundance between the different grades of periodontal health (green square in favour of health; red square in favour of pathology). The species that belonged to the core of a certain grade are coloured orange.

| No. OF OTU | GENUS | SPECIES | PERIODONTAL HEALTH GRADES |  |  |  |  |  |  |  |
| --- | --- | --- | --- | --- | --- | --- | --- | --- | --- | --- |
|  |  |  | ABUNDANCE (%) |  |  |  | DIFFERENTIAL ABUNDANCE |  |  |  |
|  |  |  | 0 | 123 | 1 | 23 | 0 vs 123 | 0 vs 1 | 0 vs 23 | 1 vs 23 |
| Otu0005 | Haemophilus | parainfluenzae | 5.43 | 3.79 | 4.42 | 3.21 |  |  |  |  |
| Otu0024 | Capnocytophaga | leadbetteri | 1.69 | 0.62 | 0.64 | 0.60 |  |  |  |  |
| Otu0037 | Oribacterium | unclassified | 0.65 | 0.38 | 0.44 | 0.33 |  |  |  |  |
| Otu0060 | Solobacterium | moorei | 0.15 | 0.40 | 0.38 | 0.42 |  |  |  |  |
| Otu0100 | Aggregatibacter | sp. HMT_512 | 0.21 | 0.10 | 0.10 | 0.09 |  |  |  |  |
| Otu0108 | Tannerella | forsythia | 0.05 | 0.17 | 0.05 | 0.29 |  |  |  |  |
| Otu0114 | Mycoplasma | faucium | 0.02 | 0.15 | 0.05 | 0.25 |  |  |  |  |
| Otu0143 | Bacteroidetes_[G-5] | bacterium_HMT_511 | 0.01 | 0.08 | 0.03 | 0.13 |  |  |  |  |
| Otu0151 | Gracilibacteria_(GN02)_[G-2] | bacterium_HMT_873 | 0.05 | 0.11 | 0.01 | 0.21 |  |  |  |  |
| Otu0181 | Cardiobacterium | hominis | 0.06 | 0.03 | 0.04 | 0.02 |  |  |  |  |
| Otu0211 | Campylobacter | concisus | 0.11 | 0.01 | 0.01 | 0.01 |  |  |  |  |
| Otu0213 | Peptostreptococcaceae_[XI][G-9] | brachy | 0.02 | 0.05 | 0.02 | 0.08 |  |  |  |  |
| Otu0228 | Peptostreptococcaceae_[XI][G-5] | bacterium_HMT_493 | 0.09 | 0.01 | 0.01 | 0.00 |  |  |  |  |
| Otu0239 | Treponema | sp. HMT_927 | 0.01 | 0.03 | 0.02 | 0.05 |  |  |  |  |
| Otu0244 | Peptostreptococcaceae_[XI][G-7] | unclassified | 0.12 | 0.01 | 0.01 | 0.02 |  |  |  |  |
| Otu0249 | Dialister | pneumosintes | 0.01 | 0.04 | 0.04 | 0.04 |  |  |  |  |
| Otu0264 | Fretibacterium | unclassified | 0.02 | 0.09 | 0.03 | 0.14 |  |  |  |  |
| Otu0307 | Bacteroidaceae_[G-1] | bacterium_HMT_272 | 0.00 | 0.02 | 0.02 | 0.01 |  |  |  |  |
| Otu0354 | Saccharibacteria_(TM7)_[G-4] | bacterium_HMT_355 | 0.04 | 0.01 | 0.00 | 0.01 |  |  |  |  |
| Otu0382 | Streptococcus | constellatus | 0.00 | 0.02 | 0.01 | 0.03 |  |  |  |  |
| Otu0439 | Simonsiella | muelleri | 0.03 | 0.03 | 0.00 | 0.05 |  |  |  |  |
| Otu0462 | Bacteroidetes_[G-3] | bacterium_HMT_280 | 0.00 | 0.02 | 0.01 | 0.03 |  |  |  |  |
| Otu0482 | Peptostreptococcaceae_[XI][G-4] | bacterium_HMT_369 | 0.00 | 0.01 | 0.01 | 0.02 |  |  |  |  |
| Otu0526 | Veillonella | dispar | 0.00 | 0.01 | 0.01 | 0.01 |  |  |  |  |
| Otu0586 | Selenomonas | dianae | 0.00 | 0.03 | 0.02 | 0.05 |  |  |  |  |
| Otu0617 | Lachnospiraceae_[G-2] | bacterium_HMT_088 | 0.02 | 0.00 | 0.00 | 0.00 |  |  |  |  |
| Otu0659 | Saccharibacteria_(TM7)_[G-1] | bacterium_HMT_869 | 0.01 | 0.00 | 0.00 | 0.00 |  |  |  |  |

The results for the global oral health scale revealed differential abundances for the different grades in 88 species (21.83% of the total OTUs), 22 of which were core species and 66 non-core (5.45% and 16.37% of the total OTUs, respectively). There were 22 species associated with oral health and 66 with some grade of global oral pathology. In the comparison of GG0 vs GG23, the differential abundances in the main global-health related OTUs (with >1% levels in GG0; log2foldchange values ranged from 1.71 to 0.88) were: *Porphyromonas pasteri, Haemophilus parainfluenzae, Veillonella parvula*, and *Capnocytophaga leadbetteri*. Those associated with high grades of global oral pathology (>1% levels in GG23; log2foldchange values ranged from −2.86 to −1.33) were: *Alloprevotella* sp. *HMT 308, Fusobacterium nucleatum_subsp._vincentii* and *Porphyromonas gingivalis* (Table 5). The relative abundances of total OTUs in dental, periodontal, global oral health grades, and analyses of differential abundance between grades are detailed in the supplementary data 2-4.

**Table 5.**
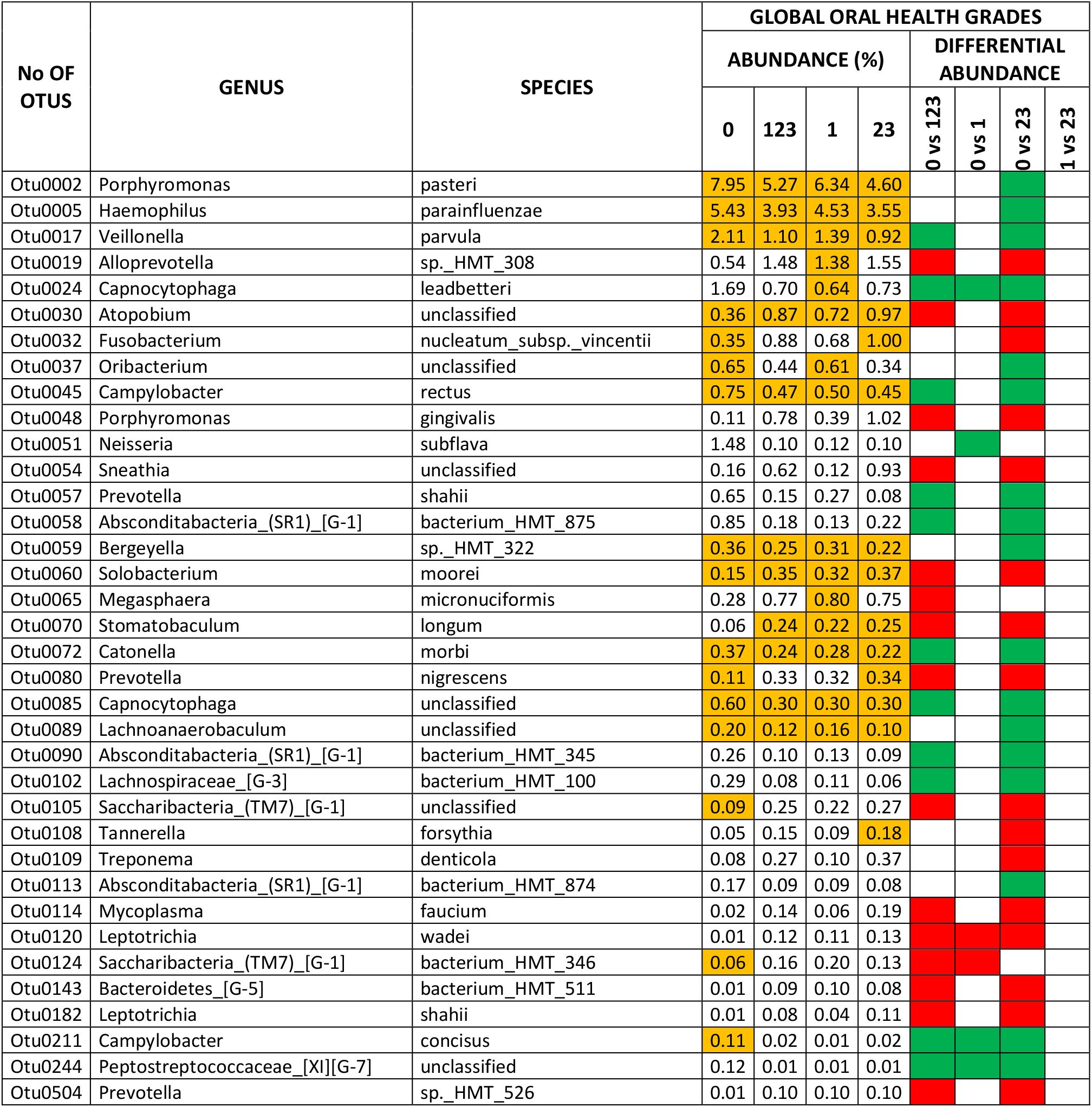
Species with abundance ≥0.1% at some grade and that presented differential abundance between the different grades of global oral health (green square in favour of health; red square in favour of pathology). The species that belonged to the core of a certain grade are coloured orange.

| No OF OTUS | GENUS | SPECIES | GLOBAL ORAL HEALTH GRADES |  |  |  |  |  |  |  |
| --- | --- | --- | --- | --- | --- | --- | --- | --- | --- | --- |
|  |  |  | ABUNDANCE (%) |  |  |  | DIFFERENTIAL ABUNDANCE |  |  |  |
|  |  |  | 0 | 123 | 1 | 23 | 0 vs 123 | 0 vs 1 | 0 vs 23 | 1 vs 23 |
| Otu0002 | Porphyromonas | pasteri | 7.95 | 5.27 | 6.34 | 4.60 |  |  |  |  |
| Otu0005 | Haemophilus | parainfluenzae | 5.43 | 3.93 | 4.53 | 3.55 |  |  |  |  |
| Otu0017 | Veillonella | parvula | 2.11 | 1.10 | 1.39 | 0.92 |  |  |  |  |
| Otu0019 | Alloprevotella | sp._HMT_308 | 0.54 | 1.48 | 1.38 | 1.55 |  |  |  |  |
| Otu0024 | Capnocytophaga | leadbetteri | 1.69 | 0.70 | 0.64 | 0.73 |  |  |  |  |
| Otu0030 | Atopobium | unclassified | 0.36 | 0.87 | 0.72 | 0.97 |  |  |  |  |
| Otu0032 | Fusobacterium | nucleatum_subsp._vincentii | 0.35 | 0.88 | 0.68 | 1.00 |  |  |  |  |
| Otu0037 | Oribacterium | unclassified | 0.65 | 0.44 | 0.61 | 0.34 |  |  |  |  |
| Otu0045 | Campylobacter | rectus | 0.75 | 0.47 | 0.50 | 0.45 |  |  |  |  |
| Otu0048 | Porphyromonas | gingivalis | 0.11 | 0.78 | 0.39 | 1.02 |  |  |  |  |
| Otu0051 | Neisseria | subflava | 1.48 | 0.10 | 0.12 | 0.10 |  |  |  |  |
| Otu0054 | Sneathia | unclassified | 0.16 | 0.62 | 0.12 | 0.93 |  |  |  |  |
| Otu0057 | Prevotella | shahii | 0.65 | 0.15 | 0.27 | 0.08 |  |  |  |  |
| Otu0058 | Absconditabacteria_(SR1)_[G-1] | bacterium_HMT_875 | 0.85 | 0.18 | 0.13 | 0.22 |  |  |  |  |
| Otu0059 | Bergeyella | sp._HMT_322 | 0.36 | 0.25 | 0.31 | 0.22 |  |  |  |  |
| Otu0060 | Solobacterium | moorei | 0.15 | 0.35 | 0.32 | 0.37 |  |  |  |  |
| Otu0065 | Megasphaera | micronuciformis | 0.28 | 0.77 | 0.80 | 0.75 |  |  |  |  |
| Otu0070 | Stomatobaculum | longum | 0.06 | 0.24 | 0.22 | 0.25 |  |  |  |  |
| Otu0072 | Catonella | morbi | 0.37 | 0.24 | 0.28 | 0.22 |  |  |  |  |
| Otu0080 | Prevotella | nigrescens | 0.11 | 0.33 | 0.32 | 0.34 |  |  |  |  |
| Otu0085 | Capnocytophaga | unclassified | 0.60 | 0.30 | 0.30 | 0.30 |  |  |  |  |
| Otu0089 | Lachnoanaerobaculum | unclassified | 0.20 | 0.12 | 0.16 | 0.10 |  |  |  |  |
| Otu0090 | Absconditabacteria_(SR1)_[G-1] | bacterium_HMT_345 | 0.26 | 0.10 | 0.13 | 0.09 |  |  |  |  |
| Otu0102 | Lachnospiraceae_[G-3] | bacterium_HMT_100 | 0.29 | 0.08 | 0.11 | 0.06 |  |  |  |  |
| Otu0105 | Saccharibacteria_(TM7)_[G-1] | unclassified | 0.09 | 0.25 | 0.22 | 0.27 |  |  |  |  |
| Otu0108 | Tannerella | forsythia | 0.05 | 0.15 | 0.09 | 0.18 |  |  |  |  |
| Otu0109 | Treponema | denticola | 0.08 | 0.27 | 0.10 | 0.37 |  |  |  |  |
| Otu0113 | Absconditabacteria_(SR1)_[G-1] | bacterium_HMT_874 | 0.17 | 0.09 | 0.09 | 0.08 |  |  |  |  |
| Otu0114 | Mycoplasma | faucium | 0.02 | 0.14 | 0.06 | 0.19 |  |  |  |  |
| Otu0120 | Leptotrichia | wadei | 0.01 | 0.12 | 0.11 | 0.13 |  |  |  |  |
| Otu0124 | Saccharibacteria_(TM7)_[G-1] | bacterium_HMT_346 | 0.06 | 0.16 | 0.20 | 0.13 |  |  |  |  |
| Otu0143 | Bacteroidetes_[G-5] | bacterium_HMT_511 | 0.01 | 0.09 | 0.10 | 0.08 |  |  |  |  |
| Otu0182 | Leptotrichia | shahii | 0.01 | 0.08 | 0.04 | 0.11 |  |  |  |  |
| Otu0211 | Campylobacter | concisus | 0.11 | 0.02 | 0.01 | 0.02 |  |  |  |  |
| Otu0244 | Peptostreptococcaceae_[XI][G-7] | unclassified | 0.12 | 0.01 | 0.01 | 0.01 |  |  |  |  |
| Otu0504 | Prevotella | sp._HMT_526 | 0.01 | 0.10 | 0.10 | 0.10 |  |  |  |  |

## DISCUSSION

The global oral health scale employed in this study was first presented by our research team in 2013, with our goal being to develop a scale that produces a single numerical value (18) to reflect a patient’s oral health status, including the presence of dental and periodontal conditions. In this study, the final dental or periodontal subscale grade corresponds to the grades assigned to at least two of the three variables analysed for each subscale. The subscale with the highest grade determined the grade for the global oral health scale (18). To the best of our knowledge, this is the first 16S-based microbiome research to determine the individualised impact of different grades of dental, periodontal and global oral disease on salivary microbiota, evaluating which of them shows the highest impact on this bacterial community.

It should be noted before our main findings are discussed that their difference to previous studies should be interpreted with caution. In particular, it is well-known that each step of the 16S rRNA gene NGS workflow may lead to pitfalls and biases that affect, for example, a study’s design and sample collection, the nucleic acid extraction and sequencing, and the bioinformatic and statistical analyses (55,56). This lack of a standardised workflow has led to uncertainty regarding the transparency, reproducibility and comparability of 16S-based microbiome research (57). In the present study, other factors that should also be taken into account are the type of saliva sample analysed (stimulated *vs* non-stimulated) (58), as well as the different clinical criteria used to define the dental and periodontal health status.

### Impact of the dental and periodontal subscales and the global oral health scale on the salivary microbiota: alpha diversity indicators and structure of the bacterial community

Various 16S-based microbiome studies on the impact of dental decay on the salivary microbiota of adults have used the DMFT index to place individuals in a healthy or a caries group (25,59,60), with the latter including subjects with active caries (DMFT ≥6) (59) or inactive tooth decay (60). In their study, Rupf et al. (61) used the Nyvad criteria to define caries activity and recruit patients for their sample who had at least three caries lesions that reached the dentine; they also used the DMFT Index and the ICDAS (61). As findings derived from these studies, the saliva samples from the healthy and caries groups generally had similar levels of richness and diversity (25,59-61). This was true whether the diseased group was composed of subjects with active (59), inactive (60) or cavitated (61) caries. In the present series, the salivary microbiota of those with the highest dental pathology grades (DG23) was significantly richer in bacteria than that of those who had a healthy dental grade (DG0); DG23 also showed a trend towards increased diversity and evenness. It should be noted that the dental health subscale incorporates variables that not only include the number of caries and their severity, but also the levels of supragingival plaque, all of which could affect the bacterial richness of the salivary community.

In most of the papers that characterise the salivary microbiome of adults with various periodontal health statuses, the classification system of the American Academy of Periodontology (62) is used to obtain diagnoses (26, 63-66). Indeed, only Chen et al. (67) have used their own diagnostic criteria, which took into account a patient’s probing pocket depth (PPD), clinical attachment loss (CAL), bleeding on probing (BOP) and gingival redness/oedema. In our study series, the periodontal health subscale was defined by levels of gingival inflammation and the number of periodontal pockets and their severity.

There is some inconsistency between the alpha-diversity results of various studies in the literature and our findings, with several of the former describing greater richness (26,67), diversity (67) and evenness (26) in the saliva samples of patients with chronic periodontitis than in those who were periodontally healthy. In our study, however, and like other authors (64), worsening periodontal health revealed a trend of increased alpha diversity, but these results were not statistically significant for any of the related parameters used in our between-grade comparisons.

Our results for the global oral health scale revealed an association between a higher grade and significant increases in the richness and diversity of the salivary microbiota. This was particularly the case for the higher grades, which are determined using a combination of dental and periodontal parameters. In line with the findings of Takeshita et al. (28), our results demonstrate the potential impact of the simultaneous presence of dental and periodontal disease on the richness and diversity of the salivary microbiota.

It has been found in various studies that the structure of the global salivary microbiota is similar in patients with good oral health (68-70). This is supported by the findings of our research, where the structure of the salivary bacterial community in the participants with oral health (grade 0) was different to that of the patients with dental, periodontal or global oral disease, especially high grades.

### Impact of the dental and periodontal subscales and the global oral health scale on the salivary microbiota: composition of the core microbiome and testing differential abundance

Numerous 16S-based microbiome studies on salivary microbiota in the literature have only conducted an analysis of the differential abundance of the taxa associated with various oral conditions (26,29,59,61,71). In this study, it is our view that it is essential to analyse salivary microbiota from a dual perspective: the prevalence of the taxa determining the core microbiome; and their differential abundance in relation to the different DGs, PGs and GGs.

The issue of taxonomic assignment is dealt with in some studies of salivary microbiota by using gender as the lowest phylogenetic level (25,26,71). The 16S rDNA sequencing-based technique, especially from Illumina platforms, lacks the necessary resolution to produce an identification of all OTUS at the species level (72,73); however, identification at, at least, this level is desirable in the oral microbiota. In our study, we have been able to verify how different species from the same genus are associated with different oral conditions, for example: *Porphyromonas catoniae* was a core species linked to dental and periodontal health, while *Porphyromonas endodontalis* was associated with dental and periodontal pathology. In relation to the differential abundance data, while *Fusobacterium periodonticum* was present in significantly higher numbers in the dentally healthy (DG0), this was only the case for those with high grades of dental pathology (DG23) in *Fusobacterium nucleatum* subsp. *vicentii*. Even, some authors have demonstrated that different strains of the same species of *Veillonella parvula* had different implications for oral health or disease (59,61). This would justify the claim of Callahan et al. (74) concerning the biological importance of conducting 16S rRNA gene-based analyses at the bacterial variant level.

In terms of prevalence, core species in the literature on the salivary microbiome have various definitions and, as a consequence, any associated findings are difficult to compare (28,59,63,64). In this study, the prevalence of grade-specific core species was less than 2.5% (≤10 OTUs), that is, they were only present in those with dental or periodontal health, or those with dental or periodontal pathology. While the abundance of the G0 specific core was 8% for dental health and 1% for periodontal health, this figure was around 2.5% for the G23 specific core in both subscales. Interestingly, about 9% (35 OTUs) of the taxa were non-specific core species; in other words, they were present in all the participants, regardless of their dental or periodontal-health grades. Moreover, most of them (about 70%) were common to both subscales, while the non-specific core’s abundance was approximately 50-57%. We have demonstrated for the first time that the non-specific core of the salivary microbiome comprises a greater number of species in higher abundances than the specific-core associated with a particular dental or periodontal condition (healthy or pathology). Interestingly, more than half of the non-specific core species in the present series were the same as those previously identified by Takeshita in ≥75% of Japanese adults (28); some of the most predominant species with an abundance ≥1% were: *Granulicatella adiacens, Haemophilus parainfluenzae, Prevotella* sp. *(P. melaninogenica), Rothia mucilaginosa, Streptococcus* sp. *(S. salivarius)*, and *Veillonella* sp. *(V. parvula)*. These results confirm that several bacterial taxa in the salivary microbiome could be present in ethnically diverse populations, regardless of their dental and periodontal health statuses.

In terms of our differential abundance findings, the number of taxa present in the salivary microbiota at significantly different levels for the subscale or global scale grades did not exceed 25%, and mainly included non-core species (5-16%); for the global oral health scale, the figure was 22%. This number of taxa was higher for the dental than the periodontal subscale (25% *versus* 7% of the total OTUs). This is the first time that this finding has been reported in the literature. If it is assumed that the salivary microbiome comprises a mix of bacterial communities originating from various sites in the oral cavity (58), our previous observations have provided evidence that the relative abundances of the most predominant bacteria in saliva are not strongly related to the oral health grade.

This corroborates the view that the dominant source of the salivary microbiome is probably the bacterial communities on the mucosal surfaces, especially those from the dorsum of the tongue (58,75,76).

When we compared our two subscales, we detected that parameters like supragingival plaque levels and the number and severity of caries had a greater differential abundance impact on the salivary microbiota than periodontal parameters such as levels of gingival inflammation and the number and severity of the periodontal pockets. Consequently, it is clear that supragingival dental conditions affect the abundance of salivary microbiota more than subgingival periodontal conditions, with the former having a greater influence on global oral health. Consequently, in line with previous authors (58), our results question the use of the salivary microbiome as an accurate indicator in the diagnosis and monitoring of caries and periodontitis, being especially evident in the latter pathology. Nevertheless, in the present series, specific bacteria were involved in different oral conditions. In our view, for a species to be evaluated as a possible bacterial biomarker associated with a given oral condition, it must be: predominant in most individuals (core species), especially in those with the oral condition with which there is an intended association; and present in high levels of abundance. Applying these criteria to our 16S-based data, the main bacteria associated with dental health (a 2-3 fold increment in relative abundance in DG0) were: *Porphyromonas pasteri, Fusobacterium periodonticum* and *Veillonella parvula*; for periodontal health (1.69-fold increase in the relative abundance in PG0), the species was *Haemophilus parainfluenzae*. All of these bacteria continued to be associated with global oral health (1.5-2-fold increase in relative abundance in GG0). The exception was *Fusobacterium periodonticum*, probably because it has been found to be more abundant in patients with periodontitis (27). These outcomes are in accordance with the findings of most 16S-based studies concerning the identification of the above-mentioned taxa as core species present in >70% of people (28,59,63,70). Other authors have, like us, also detected differential abundances of these species, which provide support for dental or periodontal health (27,59-61,67).

In our study, the main bacteria associated with high grades of dental pathology (2.5-3.5-fold increment in relative abundance in DG23) were: *Streptococcus parasanguinis* clade 411, *Alloprevotella* sp. *HMT 308, Atopobium* sp., and *Fusobacterium nucleatum_*subsp._*vincentii*. Although most of these bacteria have been described as core species in other studies (28), there is no link to dental pathology in relation to differential abundance. In the periodontal subscale, *Tannerella forsythia* was the only species associated with high periodontal pathology grades (6-fold increment in relative abundance in PG23). This bacterium has also been identified in the core microbiome in our previous studies (27,65,67). However, although it was abundant in the subgingival samples taken from participants with periodontitis (66), these levels in the salivary microbiota were not high (<0.3%) compared to those of the species mentioned previously. With respect to the other two species that compose the red complex, *Porphyromonas gingivalis* and *Treponema denticola*, although they did not present differential abundance, these taxa were ten and six times more abundant, respectively, in the highest grades of periodontal pathology than in periodontal health.

Only *Fusobacterium nucleatum_*subsp._*vincentii* continued to be the taxon associated with the highest levels of global oral pathology, with a 3-fold increase in GG23. Interestingly, although not a core species, *Porphyromonas gingivalis* was nine times more abundant for the highest global oral pathology grades, revealing its role not only in periodontal but also in dental pathology, as reported in the literature (77,78).

The main limitation of this study is clinical, due to the difficulty in recruiting patients with the highest level of periodontal pathology. This meant that grade 2 and grade 3 patients had to be placed in the same group, despite our preference to conduct an analysis for each grade individually. Although this was not an objective in the present study, another interesting avenue for future research would be to evaluate the impact that smoking has on the salivary microbiota of patients with different grades of dental, periodontal or global health.

In conclusion, the simultaneous presence of dental and periodontal pathology has a potentiating impact on the richness and diversity of the salivary microbiota. The structure of the bacterial community in oral health differs from that present in dental, periodontal or global oral disease, especially in high grades. The non-specific microbiome core comprises a greater number of species present in higher abundance than the specific core of a particular dental or periodontal condition (health or pathology). The number of taxa in the salivary microbiota with differential abundances between the DGs, PGs or GGs represents, at most, a quarter of the bacterial community and are mainly non-core species. Supragingival dental parameters condition the abundance of the microbiota more than subgingival periodontal parameters, with the former contributing more to the impact that global oral health has on the salivary microbiome.

## Data Availability

The raw sequences obtained were deposited in the SRA database under accession number PRJNA623352

## AUTHORS AND CONTRIBUTORS

Selection of patients and collection of salivary samples: MM, SF, PJJ, CC; Extraction of DNA from salivary samples: HC; Bioinformatic and statistical analysis: BCC, RIA, IT; Study of the literature: RIA, IT; Preparation and review of the manuscript: BBC, RIA, IT.

## ACKNOWLEDGEMENTS

The authors would like to thank the Genomics Unit at the Centre de Regulació Genòmica (CRG, Barcelona, Spain) for assistance with the 16S rDNA gene amplicon sequencing.

## SUPPLEMENTARY FILES

Supplementary data 1. Species belonging to the salivary core microbiome.

Supplementary data 2. The relative abundance of OTUs in dental health grades, and analyses of differential abundance between grades.

Supplementary data 3. The relative abundance of OTUs in periodontal health grades, and analyses of differential abundance between grades.

Supplementary data 4. The relative abundance of OTUs in global oral health grades, and analyses of differential abundance between grades.

